# Pharmacotherapy for Depression in Long-Term Care: A Real-World EHR Study

**DOI:** 10.64898/2026.03.13.26348347

**Authors:** Tyler M. Saumur, Katherine E. Mathers, Huda Ashraf, Brittin L. Wagner

## Abstract

**Objectives:** To evaluate rates of treatment for depression and identify resident- and facility-level predictors of pharmacotherapy among long-term care (LTC) residents in the United States.

**Design:** Retrospective, observational study.

**Setting and Participants:** Electronic health record data from 1,675,873 LTC residents in the PointClickCare Life Sciences database (January-April 2025) were reviewed and 358,425 skilled nursing facility residents with a documented depression diagnosis were identified.

**Methods:** Residents were classified as treated/untreated based on having a medication order for pharmacological depression treatment within medication classes recommended by the American Psychological Association. Descriptive analyses incorporated demographic and clinical characteristics, and multivariable logistic regression estimated odds of treatment.

**Results:** Overall, 81.7% of residents diagnosed with depression had ≥1 pharmacological depression treatment order. Selective serotonin reuptake inhibitors (59.8%) and miscellaneous antidepressants (42.3%) were the most frequently used classes. Treatment rates were similar across depression diagnoses. Higher odds of receiving treatment were observed among residents also diagnosed with vascular dementia and those with hyperlipidemia medication orders. Lower odds were noted among residents who were Black or African American, had diabetes or hyperlipidemia diagnoses, or resided in facilities located in areas with poor socioeconomic status.

**Conclusions and Implications:** Most residents with depression had at least one recommended pharmacologic therapy, although important disparities remain. Racial differences, comorbid conditions, and facility context continue to influence treatment access. These findings support the need for improved screening practices, greater attention to equity in prescribing, and strengthened clinical resources in socially vulnerable settings to enhance the quality of depression care in LTC facilities.

**Brief Summary:** Depression is common in long-term care (LTC) and is associated with poor functional and clinical outcomes, however recent treatment patterns are not well understood. Using electronic health record data from 1,675,873 U.S. LTC residents between January and April 2025, 358,425 skilled nursing facility residents were identified with a documented depression diagnosis. The use of antidepressant medication was assessed based on medication order history and was aligned with American Psychological Association recommendations. Overall, 81.7% had at least one pharmacologic treatment order for depression; selective serotonin reuptake inhibitors (59.8%) and miscellaneous antidepressants (42.3%) were most frequently used. After adjusting for covariates, lower odds of treatment were observed among Black or African American residents and among residents in facilities located in more socioeconomically vulnerable areas. These findings highlight persistent inequities in depression pharmacotherapy in LTC and support efforts to strengthen depression assessment and ensure equitable access to evidence-informed treatment across facilities.

## Introduction

Depression is a prevalent and disabling psychiatric condition among older adults in long-term care (LTC) facilities.^1^ Estimates suggest that 10% of residents experience major depressive disorder and 29**%** have significant depressive symptoms, rates far exceeding those among community-dwelling seniors.^2^ This high burden contributes to functional decline, lower quality of life, and increased mortality.^3^

Despite its impact, depression in LTC remains frequently underrecognized and undertreated, and the use of depression treatment is often poorly documented.^4,5^ For example, nearly one-third of residents without a recorded diagnosis of depression are prescribed these medications, suggesting both underdiagnosis and potential overuse.^4^ This discrepancy likely reflects antidepressant prescribing for non-depressive indications, such as anxiety or sleep disturbances, as well as incomplete or inaccurate documentation. Further, approximately one quarter of residents with a documented diagnosis of depression are not receiving antidepressant therapy, with low rates of psychotherapy in the LTC setting also reported.^4-7^ Despite early uptake of selective serotonin reuptake inhibitors (SSRIs) during their initial approval, use has decreased over time.^8^ Treatment decisions influenced by concerns about adverse drug events, drug–drug interactions, and fall risk, given that antidepressants are the psychotropic medications most strongly associated with falls among older adults.^9^

In addition, structural barriers to antidepressant use remain. Nearly half of nursing homes lack adequate psychiatric consultation, and three-quarters report difficulty accessing educational and consultative support for behavioral health concerns, contributing to the continued gap in appropriate recognition and treatment of depression.^10^ Treatment disparities extend beyond clinical need, as residents who are older, less mobile, or not White have lower odds of receiving treatment.^11^ Psychosocial interventions in nursing homes, such as the 10-week BE-ACTIV program, improve remission rates compared to usual care.^12,13^ A review of 32 trials found that psychotherapeutic and psychosocial interventions, including recreational and social engagement programs, significantly reduced depressive symptoms in residents without major cognitive impairment.^14^

These patterns raise critical questions: Who receives antidepressant therapy, what are the prevailing prescribing trends, and why do gaps persist? This study aims to quantify current treatment rates and identify resident- and facility-level predictors of depression pharmacotherapy using a large U.S. electronic health record dataset.

## Methods

### Setting and Participants

This retrospective study used EHR data from the PointClickCare Life Sciences database, which includes clinical information from over 18 million residents in U.S. LTC facilities that use PointClickCare’s EHR software including assisted living, senior living, and skilled nursing facilities (SNFs). The EHR includes deidentified and expert-determined data such as resident demographics, Minimum Data Set clinical and functional assessments, medication and vaccination records, and related data elements. Data is gathered from residents with facility-obtained consent and covered by business associate agreements with LTC facilities. Exemption from ethics committee oversight was granted by the University of Toronto and University of Alberta Research Ethics Boards.

Between January 1 and April 30, 2025, the dataset included 1,675,873 individuals, of whom 1,251,460 were SNF residents. Among these, 358,425 residents had a documented diagnosis of depression. Residents were included if they had at least one day of residence in a LTC or SNF facility during the study period, and at least one active diagnosis for depression during that time, including Major Depressive Disorder (International Classification of Diseases [ICD]-10 codes F33, F33.0, F33.1, F33.2, F33.3, F33.4, F33.40, F33.41, F33.42), Unspecified Depression (F32.A), and Other Depression (F33.8). The dataset included demographic details, comorbidities, prescribed and administered medications, and facility characteristics.

### Exposures and Variables of Interest

Residents were classified as treated or not treated, based on having a medication order for pharmacological depression treatment within medication classes consistent with the American Psychological Association (APA) guidelines for depression management.^15^ Medication classes included SSRIs, serotonin-norepinephrine reuptake inhibitors, serotonin modulators and stimulators, tricyclic antidepressants, norepinephrine-dopamine reuptake inhibitors, and monoamine oxidase inhibitors.

Demographic variables included age, sex, race/ethnicity, insurance type, and facility region. Days in facility (DIF) was calculated as the number of days between a resident’s admission and discharge dates. For residents in facility at the end of the study period, 30 April 2025 was used as the discharge date. Consecutive visits were combined when separated by gaps of seven days or fewer, treating them as a single continuous stay for DIF calculation. Clinical variables captured comorbidities commonly associated with treatment variation, such as diabetes and hyperlipidemia. Facility-level factors, including geographic location and resident census, were also evaluated.

### Data Analysis

Data were extracted using SQL and analyzed in Python. Descriptive statistics summarized resident demographics and clinical characteristics, with means and SDs reported for continuous variables and counts (%). A multivariable logistic regression model estimated the odds of receiving pharmacological therapy, adjusting for demographic, clinical, and facility characteristics, including comorbid chronic conditions.

## Results

### Demographic and Clinical Characteristics

There were 358,425 residents with depression who met the eligibility criteria (Table 1). Of those eligible residents, 81.7% had orders for depression medication. Across this study population, the mean (SD) age of residents was 74 (12.3) years. The majority of residents were female (62.3%) and White (72.5%). Facilities were mostly located in Southern (38.9%) or Midwestern (27.6%) U.S., with an even distribution of admitting payer types across Managed Care, Medicare FFS, and Medicaid.

**Table 1.**
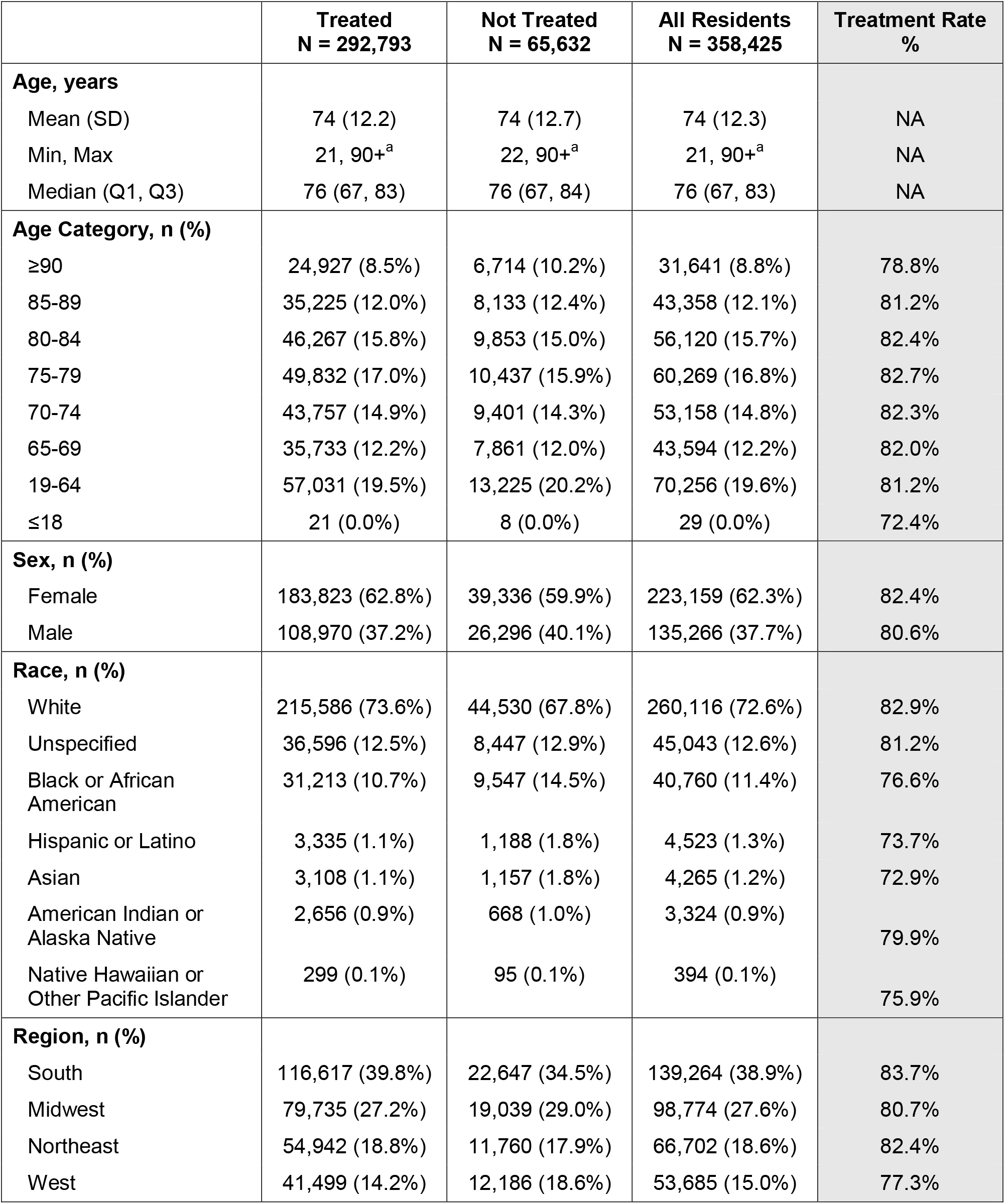
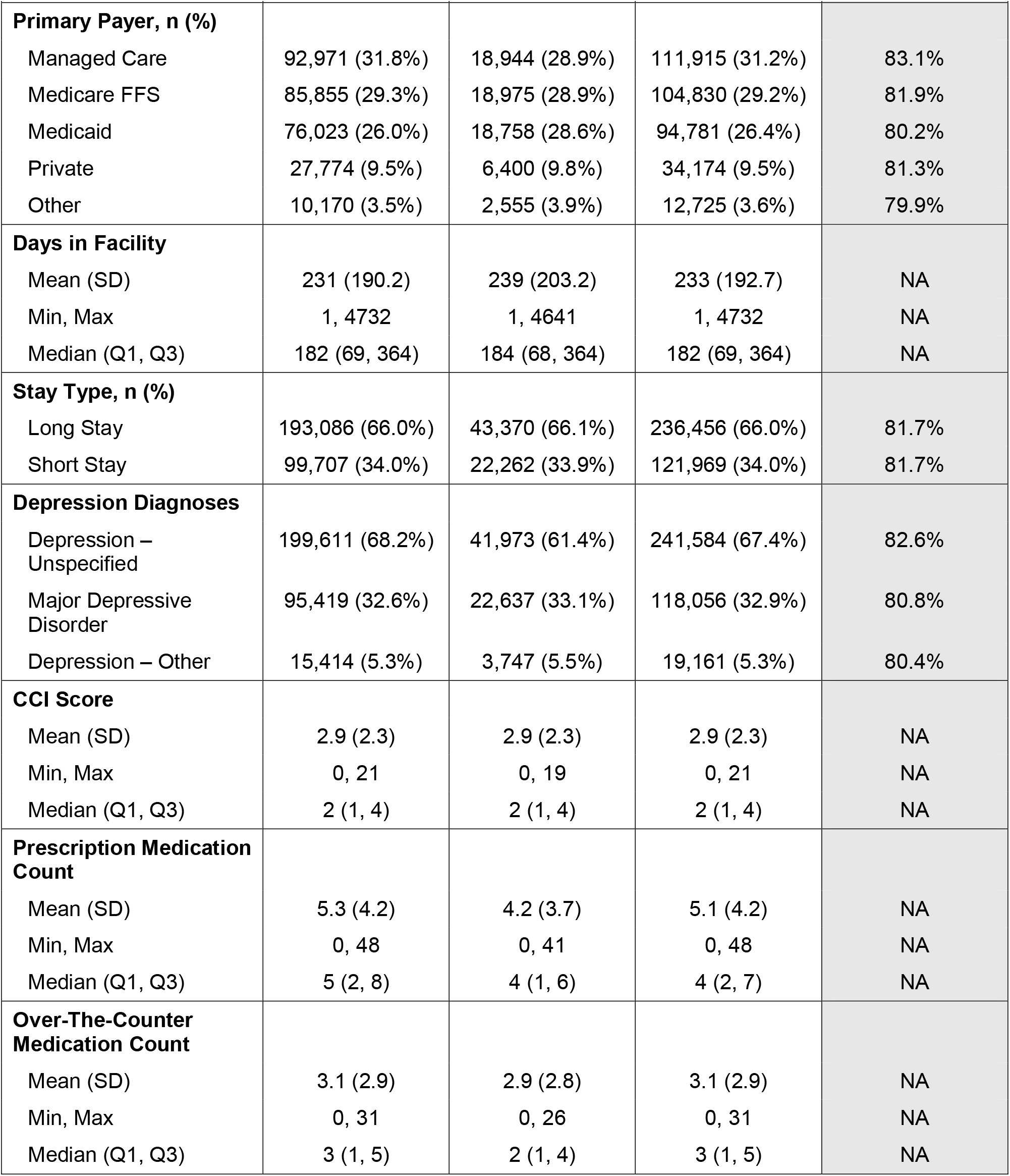
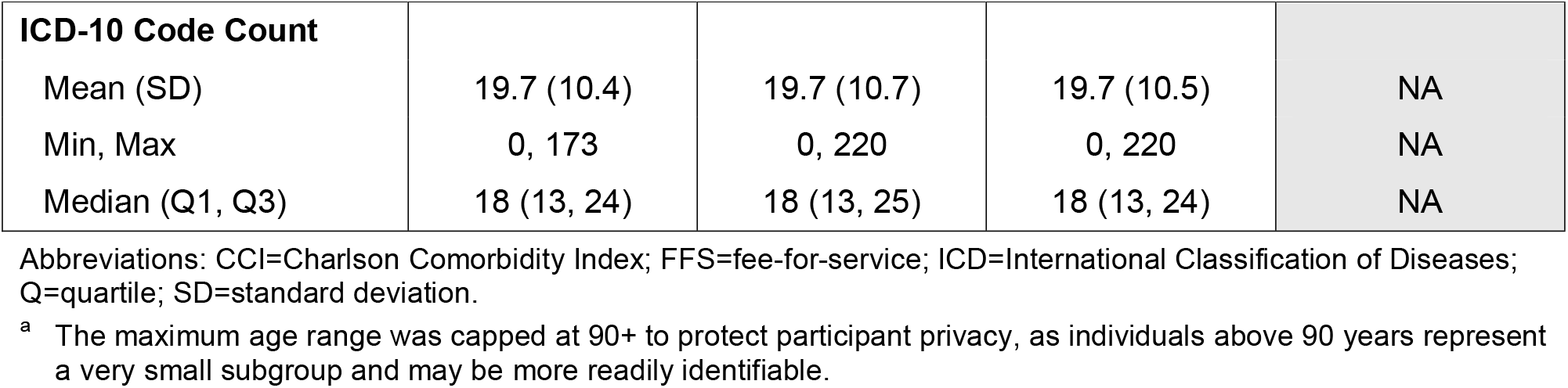
Demographic and Clinical Characteristics Across Exposure Groups.

Approximately two-thirds of residents were diagnosed with unspecified depression and some residents were diagnosed with more than one depressive condition (Table 1). The Charlson Comorbidity Index and ICD-10 code count were similar between those treated and not treated, with means of 2.9 and 19.7, respectively among all residents. With respect to medication, treated residents were taking approximately 1 more prescription medication compared to untreated residents, with similar numbers of over-the-counter medications. The most common depression medications were SSRIs (59.8%) and miscellaneous antidepressants (42.3%), which included medications like vortioxetine, mirtazapine, and trazodone (Table 2).

**Table 2.** Treatment Rates by Medication Class.

|  | **Treated**<br>**N = 292,793** |
| --- | --- |
| **Medication Class, n (%)** |  |
| Selective Serotonin Reuptake Inhibitors | 175,183 (59.8%) |
| Miscellaneous Antidepressants | 123,758 (42.3%) |
| Serotonin-Norepinephrine Reuptake Inhibitors | 65,169 (22.3%) |
| Norepinephrine-Dopamine Reuptake Inhibitors | 23,705 (8.1%) |
| Tricyclic Antidepressants | 9,683 (3.3%) |
| Serotonin Modulators and Stimulators | 1,163 (0.4%) |
| Monoamine Oxidase Inhibitors | 50 (0.0%) |

### Treatment Rates and Odds Ratios

Treatment rates were similar across all depression diagnoses. Odds ratios were generated following a multivariate logistic regression model, which comprised of 67 demographic and clinical variables (Supplementary Table 1). Odds ratios ranged from 0.71 to 2.10, with the 5 highest and lowest odds ratios displayed in Figure 1. Specific depression diagnoses had the highest odds associated with receiving depression medication, with diagnoses of Unspecified Depression and Major Depressive Disorder being most predictive of treatment. Among the variables with high treatment odds were factors associated with other (non-depression) chronic conditions; having a diagnosis of vascular dementia as well as taking any hyperlipidemia medication both had high odds of receiving treatment for depression. The variable with the lowest odds of receiving treatment was Black or African American race (odds ratio = 0.71), with diagnoses of diabetes or hyperlipidemia, and in a facility with a high social vulnerability index with the next lowest odds of receiving treatment.

**Figure 1.**
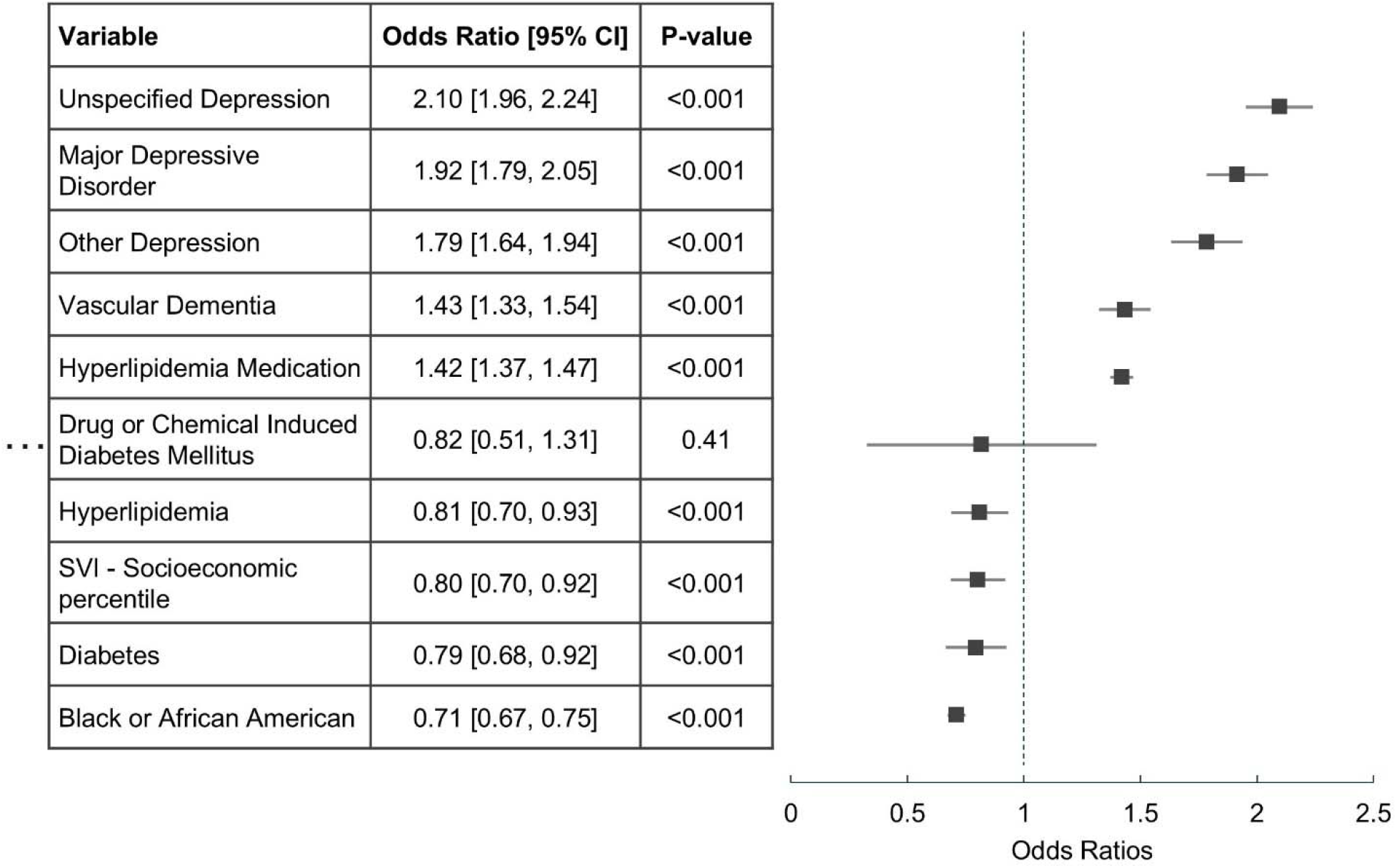
Five Highest and Lowest Odds Ratios. Abbreviations: SVI=Social Vulnerability Index; CI=confidence interval

## Discussion

This study explored rates of pharmacological treatment and factors associated with pharmacologic depression care in a large U.S. electronic health record dataset. Overall, 81.7% of residents diagnosed with depression received treatment within medication classes recommended by the American Psychological Association, with SSRIs and miscellaneous antidepressants as the most commonly used medications. Those with vascular dementia and taking hyperlipidemia medication had higher odds of receiving treatment, whereas those who were Black or African American, diabetic, in a facility in a poor socioeconomic area, or had hyperlipidemia were at decreased odds.

The proportion of the population receiving medication for depression in the present study aligns with previous work in nursing facilities from approximately 20 years ago.^4,16^ While awareness and attention to depression and depression treatment options have increased over the years, one might expect that treatment rates would also increase in tandem. In the LTC setting however, factors such as polypharmacy and comorbidities that may take higher priority in their care journey may ultimately limit the number of individuals with depression being treated. Furthermore, trends such as race and socioeconomic status of areas continue to play a role on whether or not individuals receive depression treatment. Socioeconomic status was based on a number of Census variables including below 150% poverty, unemployment, housing cost burden, no high school diploma, and no health insurance.^17^ Those in facilities in the lower socioeconomic percentiles had significantly low odds of receiving depression treatment.^18^ This may be due to staffing disparities and limited resources in areas with poorer socioeconomic status. The present study also found that Black or African American residents had the lowest odds of receiving depression treatment. Key barriers to seeking treatment in this population include stigma and a lack of confidence and trust in mental health care.^19,20^ While approaches to combat these barriers such as psychoeducation and eliminating microaggressions during therapy have been suggested,^21^ our findings appear to suggest that these disparities in treatment are still present.

The results of the study also showed that those taking hyperlipidemia medication had an increased odds of receiving depression treatment, despite those with a diagnosis of hyperlipidemia having a decreased odds of receiving depression treatment. These findings may appear to be at odds with one another; however, we need to consider that those taking daily hyperlipidemia medications i.e., statins, fibrates etc. are also more likely to get refills for other prescriptions that they are taking, such as depression medications. It may be that those untreated for depression, may also be the same group of individuals diagnosed, but not being treated for hyperlipidemia. Indeed, we showed that are 25% of individuals diagnosed with hyperlipidemia are untreated.^22^ Understanding the interplay between the comorbidities and their impact on disease-specific care is needed.

Despite these insights, the study has several limitations. Firstly, non-medication treatments for depression such as cognitive behavioral therapy and interpersonal psychotherapy were not considered due to the pharmacological scope of this study. In addition, other factors not included in this study such as facility operations, staffing numbers etc. may also influence treatment rates and patterns. Furthermore, the cross-sectional nature precludes causal inference, and unmeasured confounding variables could have influenced the observed associations. Additionally, the generalizability of the findings is limited to LTC populations and similar healthcare systems and does not necessarily reflect treatment patterns in the community. Nevertheless, the study findings are notable as they represent a substantial proportion of the LTC population in the United States diagnosed with depression.

## Conclusions and Implications

The findings of this study reveal persistent gaps in the delivery of pharmacological treatment among LTC residents with depression, with notable disparities affecting residents who are Black or African American, those with diabetes or hyperlipidemia, and individuals living in socially vulnerable facilities. Although overall treatment rates were high, these inequities indicate that access to recommended therapies remains uneven across clinical and demographic groups. The increased likelihood of treatment among residents with vascular dementia and those already receiving chronic disease medications suggests that greater medical oversight may improve identification and management of depression. The use of antidepressants to treat depressive symptoms or behavioral symptoms related to vascular dementia remains unclear due to low uptake of indications associated with prescriptions being provided.

From a practice and policy standpoint, these results underscore the need for targeted strategies to strengthen depression screening, support equitable prescribing, and enhance access to mental health expertise within LTC settings. Due to the safety risks associated with depression pharmacotherapy, appropriate safety management and monitoring are vital to ensuring safe prescribing in this population. Improving provider education and integrating decision-support tools may further promote consistent, evidence-based care. Future research should focus on identifying the drivers of treatment disparities and evaluating interventions that advance equitable and effective depression management for this vulnerable population.

## Data Availability

Data produced in the present study are available upon reasonable request to the authors.

## Acknowledgements

We would like to thank Jody Long and Dr. Steve Buslovich for their collaboration and their clinical input provided for this manuscript. This research was supported by PointClickCare Life Sciences and McMaster University. McMaster University Library provided journal article access.

## Supplementary Tables

**Supplementary Table 1.**
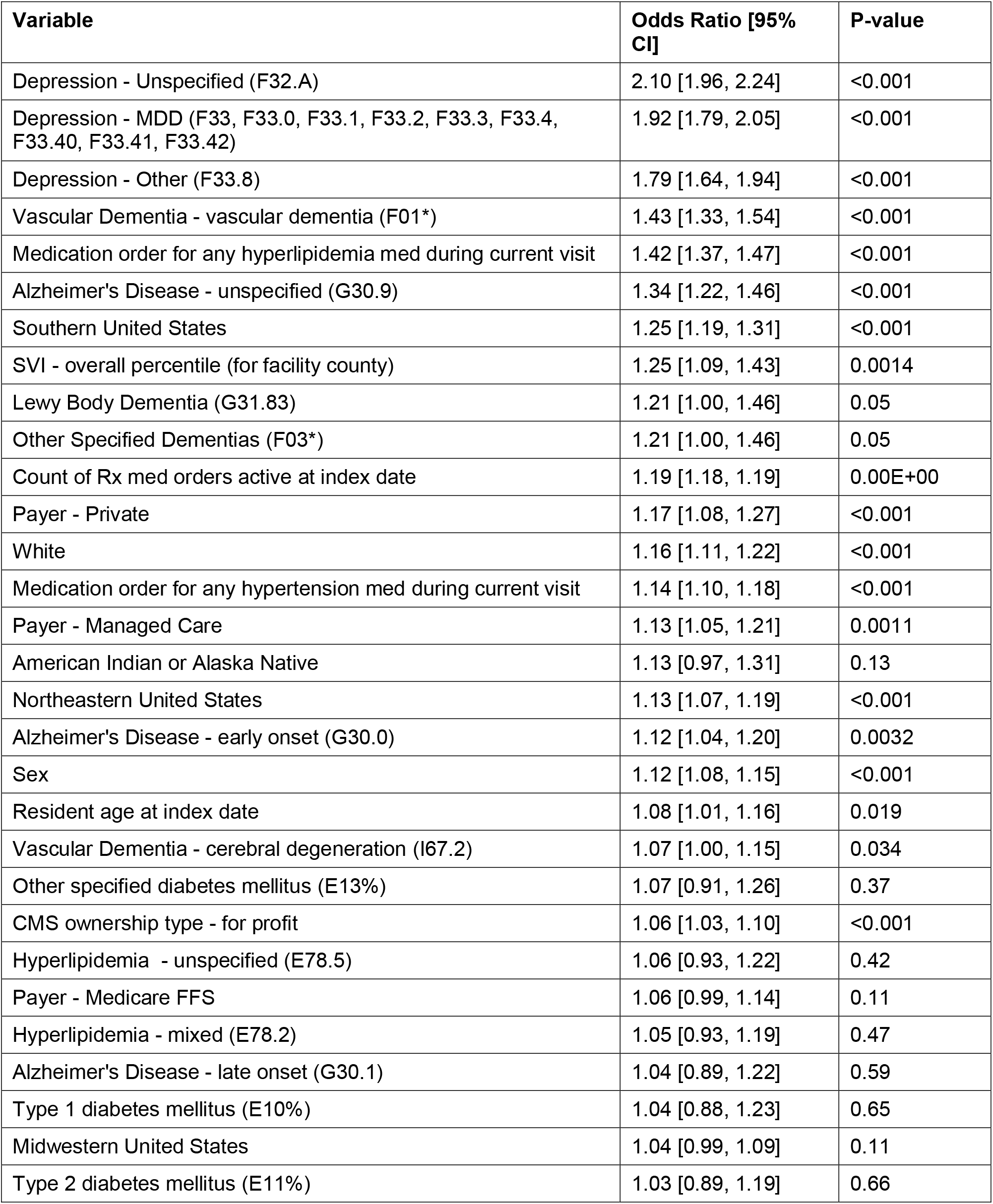
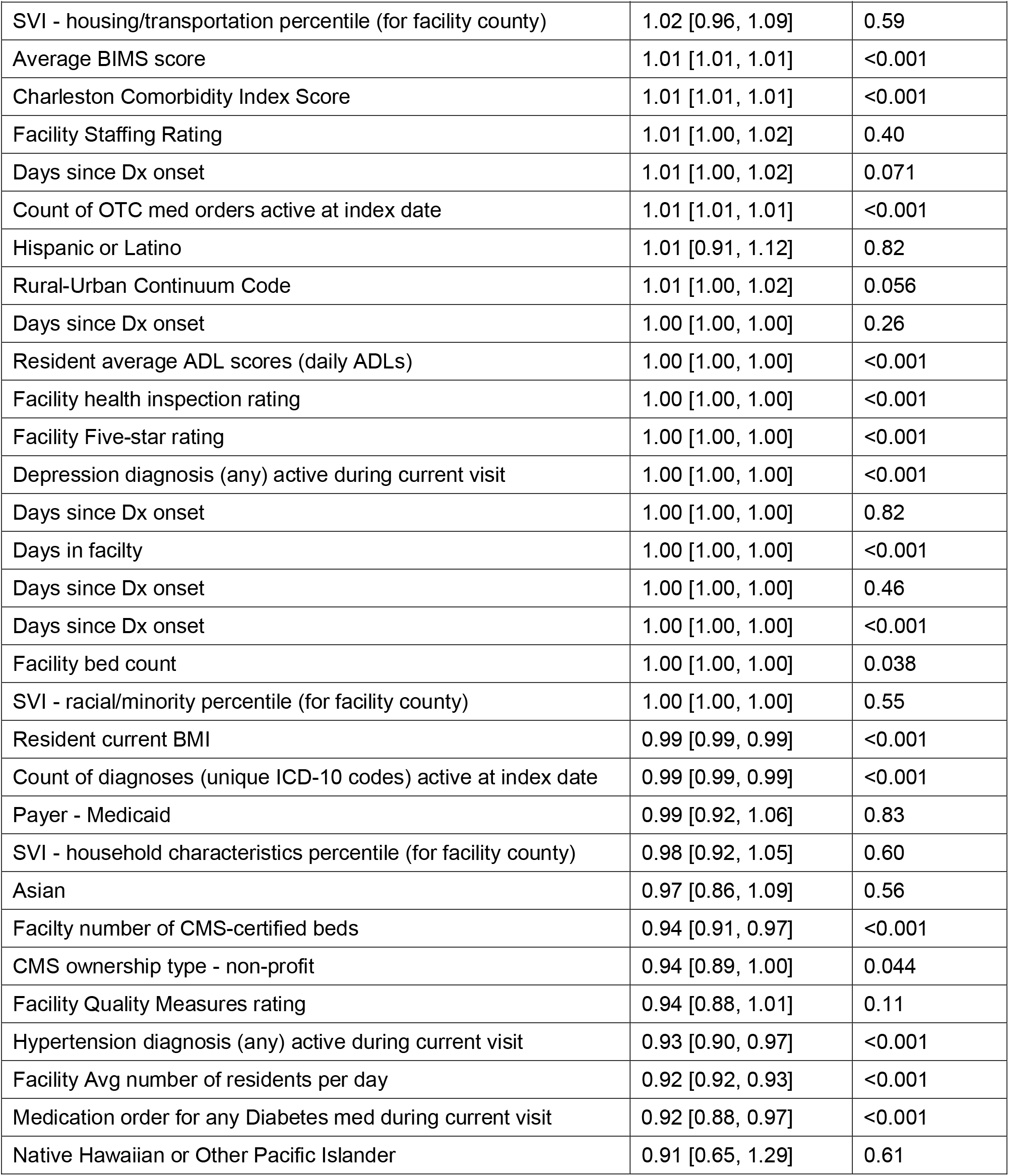
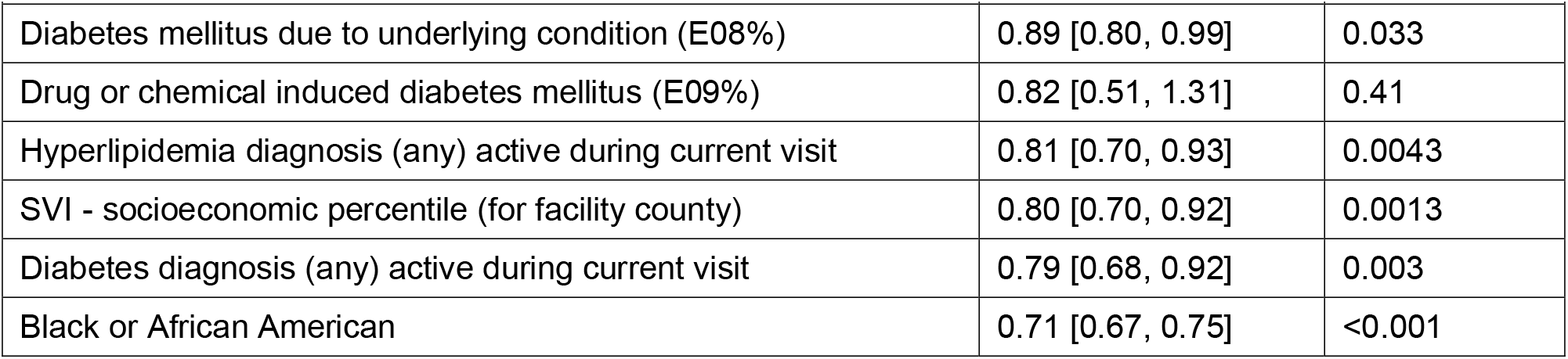
Complete List of Variables and Odds Ratios in the Multivariate Logistic Regression Model.

